# Somatic RIT1 indels identified in arteriovenous malformations hyperactivate RAS-MAPK signaling and are amenable to MEK inhibition

**DOI:** 10.1101/2023.11.13.23298448

**Authors:** Friedrich G. Kapp, Farhad Bazgir, Nagi Mahammadzade, Erik Vassella, Yvonne Döring, Annegret Holm, Axel Karow, Caroline Seebauer, Natascha Platz Batista da Silva, Walter A. Wohlgemuth, Pia Kröning, Charlotte M. Niemeyer, Denny Schanze, Martin Zenker, Whitney Eng, Mohammad R. Ahmadian, Iris Baumgartner, Jochen Rössler

## Abstract

Arteriovenous malformations (AVM) are benign vascular anomalies prone to pain, bleeding, and progressive growth. Treatment is often difficult and relapse after therapy is common. AVM are mainly caused by somatic mosaicism with pathogenic variants of the RAS-MAPK pathway. However, a causative variant is not identified in all patients. Using ultra-deep next generation sequencing we identified novel somatic *RIT1* indel variants in lesional tissue of three AVM patients. *RIT1* – not previously implicated in AVM development – encodes a RAS-like protein that can modulate RAS-MAPK signaling. For biochemical characterization, we expressed *RIT1* variants in HEK293T cells, which led to a strong increase in ERK1/2 phosphorylation. Endothelial-specific mosaic overexpression of the *RIT1* indels in zebrafish embryos induced AVM formation, highlighting the functional importance in vascular development. Both ERK1/2 hyperactivation *in vitro* and AVM formation *in vivo* could be suppressed by pharmacological MEK inhibition. Targeted treatment with the MEK inhibitor trametinib led to a significant decrease in bleeding episodes and AVM size in one patient. Our findings expand the genetic spectrum of AVM by identifying *RIT1* as a novel gene involved in AVM formation and pave the way for targeted treatment and clinical trials in patients with AVM.

## Introduction

Vascular anomalies are classified according to the Classification of the International Society for the Study of Vascular Anomalies (ISSVA) and are subdivided into vascular tumors and vascular malformations ^1,2^. While vascular tumors show increased cell proliferation, vascular malformations are thought to represent mainly non-proliferative lesions that originate from errors in vascular development. Most vascular malformations are caused by a somatic mosaic mutation in the affected tissue. Activation of the PI3K-AKT-mTOR pathway are thought to predominate in slow-flow malformations such as venous and lymphatic malformations ^3–5^, whereas variants activating the RAS-MAPK pathway are typically associated with fast-flow malformations such as AVM ^6–9^.

Extracranial AVM can occur anywhere in the body, most often in the soft tissue of extremities as well as the head and neck ^10,11^. AVM may become symptomatic with swelling, pain, pulsations, and bleeding. AVM located in the face may lead to major disfigurement and life-threatening complications ^12^. Almost all AVM progress over time ^13,14^, which may lead to tissue necrosis, bleeding complications, and hyperdynamic heart failure. Treatment is mainly interventional (embolization) and/or surgical resection; however, invasive therapies can activate the lesion and often lead to relapse ^14^. However, the exact pathomechanism of AVM development and progression are poorly understood. Taken together, AVM belongs to the most aggressive vascular anomalies and are often difficult to treat, highlighting the need for novel treatment strategies.

In this project, we identified novel mosaic indel variants in *RIT1* that were found in the lesional tissue of three patients with extracranial AVM. RIT1 acts as modulator of the RAS-MAPK pathway and has so far not been implicated in the development of AVM. We characterized these variants by assessing their effect on ERK phosphorylation *in vitro*, on vascular development *in vivo* in a zebrafish model, and the response to MEK inhibition. We further present data on the off-label use of trametinib in one patient.

## Results

### Novel *RIT1* indel variants identified in AVM tissue from three patients

Patient 1 (P1) was a girl who is in her early childhood with an AVM of the right face. A capillary anomaly and swelling of the right cheek were noticed at birth. A diagnosis of an infantile hemangioma was initially made at an external hospital and propranolol was initiated. The lesion did not respond to this therapy and a first episode of epistaxis occurred after 5 months, eventually leading to the diagnosis of a facial AVM on magnetic resonance angiography (MRA) (Fig 1A and S1A). Following this bleeding event, a first catheter embolization with Onyx was performed. Two additional embolizations were also performed, the last intervention of which was combined with bleomycin electro-sclerotherapy^15^. The lesion did not respond to either therapy and the AVM progressed with intermittent life-threatening bleeding episodes. Due to progressive symptoms, the patient was then treated with extensive Onyx embolization of the AVM, and a biopsy was obtained for genetic analysis. These interventions, including the removal of a molar that was rooted within the AVM, alleviated the symptoms only slightly. Due to the nature and course of the disease, a hemimaxillectomy was considered. However, infiltration of the AVM into the orbit made a cure by this very invasive approach seem unlikely. We thus initiated off-label treatment with thalidomide (25 mg per day). Under this treatment – and after the last extensive embolization – the severity of bleeding episodes decreased over the next months before deteriorating again. Genetic testing of the biopsy of affected tissue identified a *RIT1* indel variant (c.246_248delinsCCCTCT p.T83delinsPL (referred to as *RIT1^P^*^1^, hereafter)), with a variant allele frequency (VAF) of 3.3%.

**Fig 1.**
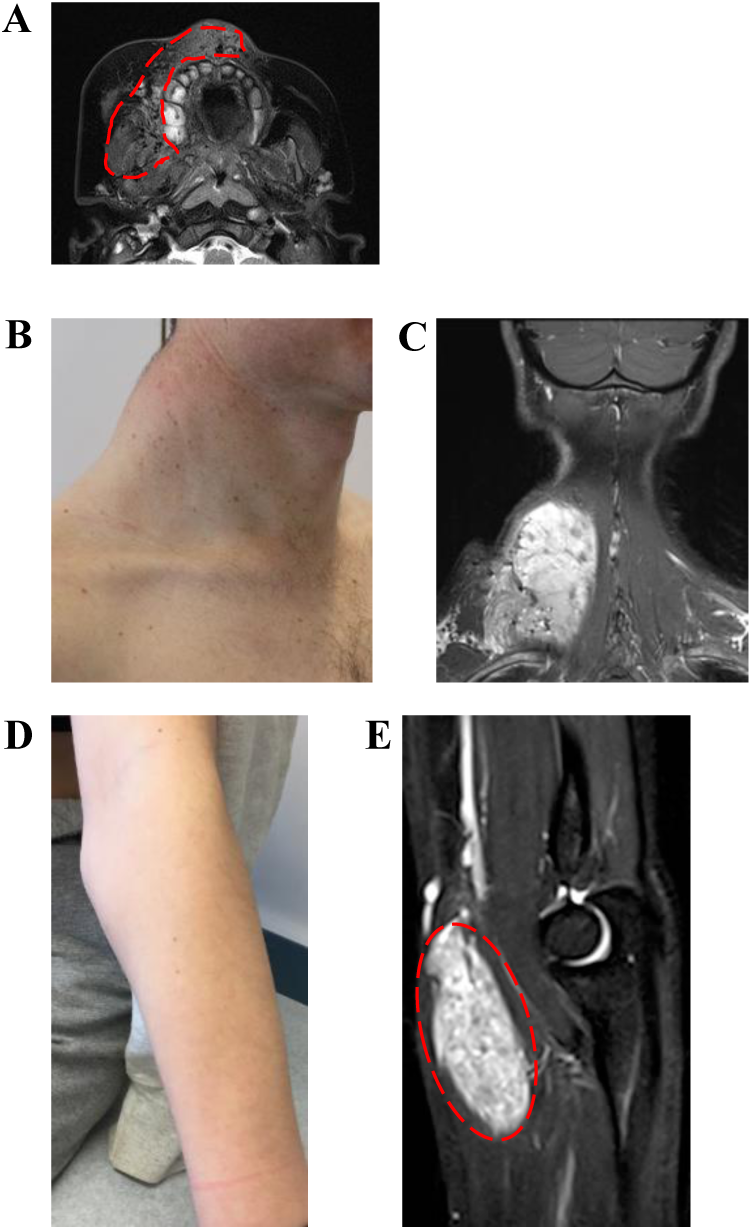
Three patients with somatic RIT1 variants identified in AVM tissue. A. MRI of P1 at the age of 4 months, the image of a transversal T2 TSE sequence, in which an AVM could be detected; the extent of the lesion is labelled with red dashed line indicating soft tissue edema and flow-voids. B. Patient P2 shows a prominent mass of the left cervical / nuchal area. C. MR imaging (coronal T2 sequence with fat saturation), which shows the hyperintense isolated intramuscular lesion, flow-voids seen within the lesion representing high AVM flow, and disfiguring overgrowth. D. Clinical aspect of the Patient P3 with swelling on the left forearm, close to the medial side of the elbow. E. T2W sagittal images demonstrating a well-defined fusiform shaped hyperintense lesion involving the flexor muscle (pronator teres) of the forearm. Flow voids (red dashed line) are seen within the lesion representing arterial blood vessels.

Patient 2 (P2) was a man in his 40s with a first episode of neck pain at the age of 30-35 years. A continuously growing and pulsating vascular lesion was detected (Fig 1B). MRA showed an isolated intramuscular AVM connected to the subclavian and the thyrocervical trunk on the right side with disfiguring diffuse muscle involvement including the splenius capitis muscle (Fig 1C and S1B). Therapy with sirolimus was initiated but had to be discontinued due to suppurative osteomyelitis of the jaw within 3 weeks. Later, the patient received three direct intraarterial ethanol embolizations at monthly intervals without success. Although the initially dominating nidus of the AVM was completely shut down, there was a massive proliferation of microfistular AV shunts and an increase in tissue volume as a result. One year after embolization, debulking surgery was performed after the situation had stabilized. Histopathology showed typical findings of a diffuse intramuscular microfistular AVM. Ten months later, progression of the AVM was noted again and a combined approach with Onyx embolization and gross total resection was performed. Since then, the patient has been without complaints with stable disease and minimal radiological residuum. A *RIT1* indel variant was identified in the resected tissue (c.242_248delinsTCCCTCT p.E81_T83delinsVPL (referred to as *RIT1^P^*^2^, hereafter) with a VAF of 6.0%.

P3 was a girl in her teens, who presented with a persistent prominence in the left forearm that was first noted one year before (Fig 1D). At the time of the initial presentation, there was no associated pain, no functional deficit, no overlying skin changes, and only minimal swelling. An initial ultrasound was notable for a 5.4 cm x 1.1 cm x 4.7 cm intramuscular mass in the left forearm with diffuse internal vascularity seen on Doppler examination. An MRI of the lesion was notable for a solid enhancing mass in the left pronator teres muscle with imaging findings consistent with a solid neoplasm (Fig 1E). She underwent an IR-guided biopsy of the lesion. Histopathology was consistent with an intramuscular fast-flow vascular anomaly. She was followed for the next two years and had progressive growth of the lesion associated with pain.

Given the worsening of her symptoms, she underwent resection of the lesion. There were no complications, and she has had minimal pain since. In the resected tissue, a *RIT1* indel variant was identified (c.229delinsTTGGATACAA p.A77delinsLDTT (referred to as *RIT1^P^*^3^, hereafter) with a VAF of 13%.

### RIT1 indel-induced ERK hyperphosphorylation can be reversed by pharmacological inhibition of MEK but not SHP2

All three indel variants are located close to the switch 2 domain of the RIT1 protein, a region that also harbors germline missense variants commonly associated with Noonan syndrome (Fig 2A and B). To investigate the impact of these novel mutations on activation of the RAS pathway, we assessed ERK phosphorylation by Western blotting after the expression of RIT1 in the HEK293T cells. We expressed RIT1^P1^, RIT1^P2^, RIT1^P3^, RIT1 wildtype (RIT1^wt^), and two recurrent RIT1 mutations found in Noonan syndrome (p.F82L and p.M90I). All three novel RIT1 indels led to a significant increase in ERK phosphorylation, while overexpression of the two Noonan syndrome-associated RIT1 mutations only induced a modest ERK hyperphosphorylation (Fig 2C, D).

**Fig 2.**
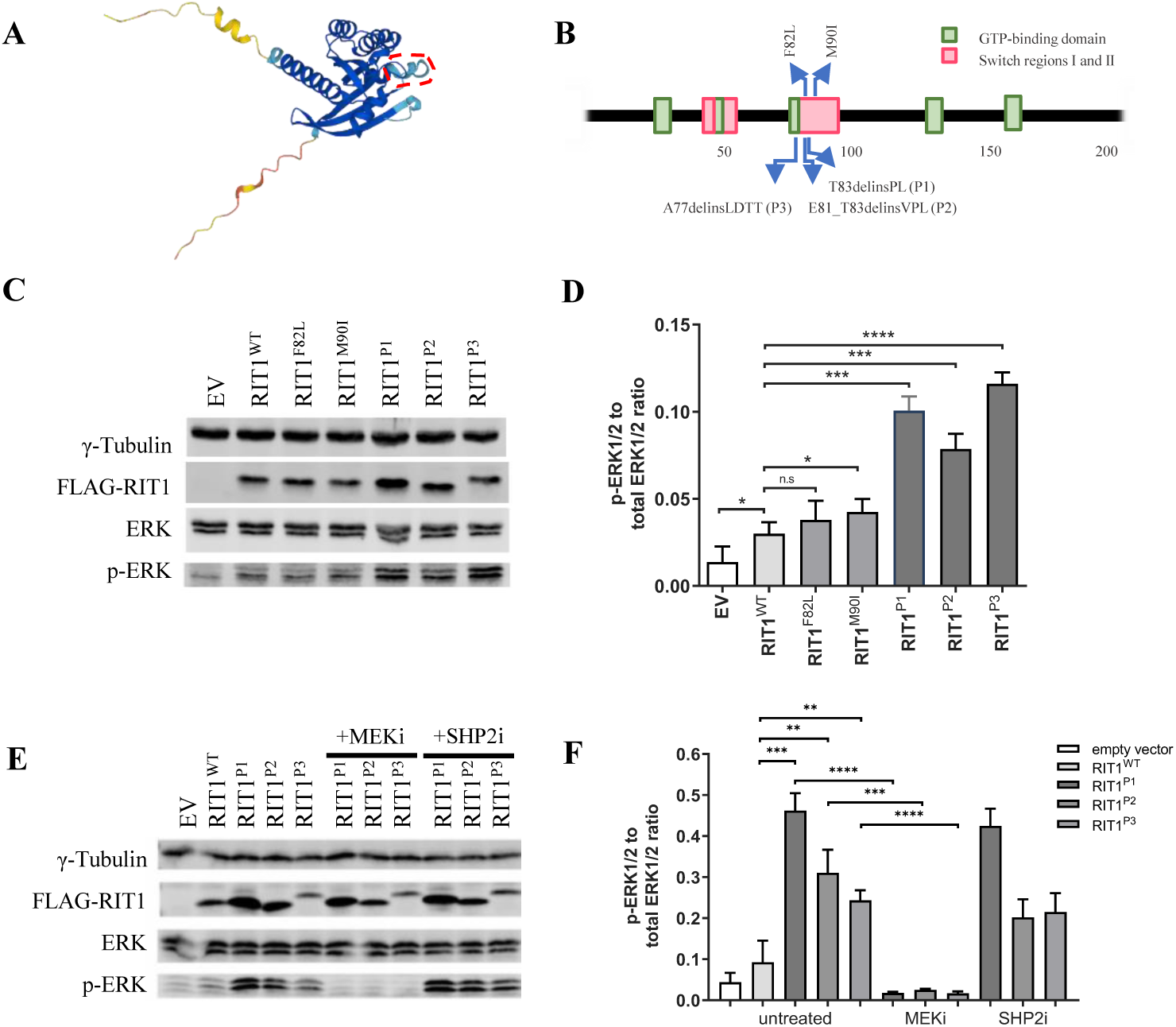
ERK phosphorylation after expression of RIT1 variants *in vitro* in HEK293T cells. A. Protein structure of RIT1 predicted by AlphaFold ^24,25^, accessed through ensemble.org. The area labelled by the dashed red line indicates the switch 2 domain. B. Schematic drawing of *RIT1* functional domains of human RIT1 protein (green boxes = GTP-binding regions; red boxes = switch domain 1 and 2; blue arrows (upward) = two mutations typically found in Noonan syndrome; blue arrows (downward) = mutations identified in P1-P3. C. Western blot after expression of *RIT1* variants to assess RAS-MAPK pathway activation. Gamma tubulin served as loading control, FLAG-RIT1 confirms the expression of the construct, total ERK levels serve as a control to exclude the differential expression of ERK, and p-ERK measures the level of phosphorylate of ERK as a marker of RAS pathway activation. D. Quantification of the ERK phosphorylation was measured in a total of three western blots (n=3). Student’s t-test, one-tailed. *P(WT vs P1) = 0.0346, P(WT vs F82L) = 0.1772, *P(WT vs M90I) = 0.0489, ***P(WT vs P1) = 0.0002, ***P(WT vs P2) = 0.0010, ***P(WT vs P3) = 0.00004. Data are presented as mean ± SD. EV = empty vector. E. Western blot after expression of *RIT1* variants and with or without treatment using a MEK inhibitor or SHP2 inhibitor. The same parameters were assessed as in panel D. F. Quantification of the ERK phosphorylation was measured in a total of three western blots (n=3). Student’s t-test, one-tailed. Untreated: ***P(WT vs P1) = 0.0003, **P(WT vs P2) = 0.0040, **P(WT vs P3) = 0.0055, MEKi: ****P(P1) = 0.00003, ***P(P2) = 0.0005, ****P(P3) = 0.0001. Data are presented as mean ± SD.

Since RAS proteins act downstream of SHP2 and upstream of MEK and ERK in the RAS-MAPK signaling pathway, we hypothesized that RIT1-induced ERK hyperphosphorylation exhibits a differential response to treatment with SHP2 and MEK inhibition (Fig S2). Indeed, treatment of HEK293T cells with the SHP2 inhibitor SHP099 showed no effects on ERK phosphorylation. In contrast, MEK inhibition with PD0325901 reversed ERK phosphorylation close to baseline levels (Fig 2E, F).

### *RIT1* indel variants lead to the formation of AVM-like lesions in zebrafish embryos

Having shown previously that the *RIT1* indels identified in AVM patients induced strong activation of the RAS-MAPK pathway, we next assessed whether their expression can lead to aberrant vascular development in the tail vasculature of zebrafish embryos. The zebrafish is an established model for the study of vascular development ^16^ that has also been applied to translational research ^8,17,18^. To this end, we used plasmids, which contain the transcriptional upstream activating sequence (UAS) that controls the expression of wildtype or variant *RIT1* linked to GFP via the self-cleaving peptide P2A. Activation of the UAS element and thus expression of RIT1 is dependent on the presence of the transcription factor Gal4. These plasmids were injected in the one-cell stage of *Tg(fli1a:Gal4; UAS:RFP)* embryos. While the plasmid integrated randomly into the DNA of cells of the zebrafish embryo, expression of RIT1-P2A-GFP was limited to endothelial cells that expressed Gal4 under the control of the endothelial fli1a promoter (Fig 3A)^18^. Using this approach, we observed AVM-like lesions in the zebrafish embryo tail. These lesions were characterized by aberrant connections between the dorsal aorta and the caudal vein. The most severe phenotypes exhibited a fusion of these arterial and venous vessels (Fig 3B-F). In some embryos, the vessels directly downstream of the AVM-like lesions were dilated, while the further distal part of the tail vasculature was hypoplastic (Video S1). A significantly higher rate of AVM-like lesions at 48 hours post fertilization (hpf) was observed in embryos expressing *RIT1^P^*^1^*^-P^*^3^ indels compared to *RIT1^wt^* (66- 75% vs 24%, Fig 3G). Next, we treated injected embryos with 100 nM trametinib during early development. This early treatment significantly reduced the formation of AVM-like lesions (31-39%, Fig 4A and B), thereby supporting the assumption that AVM formation is critically dependent on hyperactivation of the RAS-MAPK pathway. To mimic a targeted treatment more closely in patients, we next treated zebrafish embryos with established AVM-like lesions after injection of *RIT1* indels with 100 nM trametinib from 48 hpf onwards and compared growth of the lesions in treated and untreated embryos over the following two days. DMSO-treated embryos in the control group showed a relative increase in the size of the lesion to 117.5%, while the size of the lesions in trametinib-treated embryos decreased to 83.4%; the difference between treated and untreated embryos was significant (Fig 4C and D).

**Fig 3.**
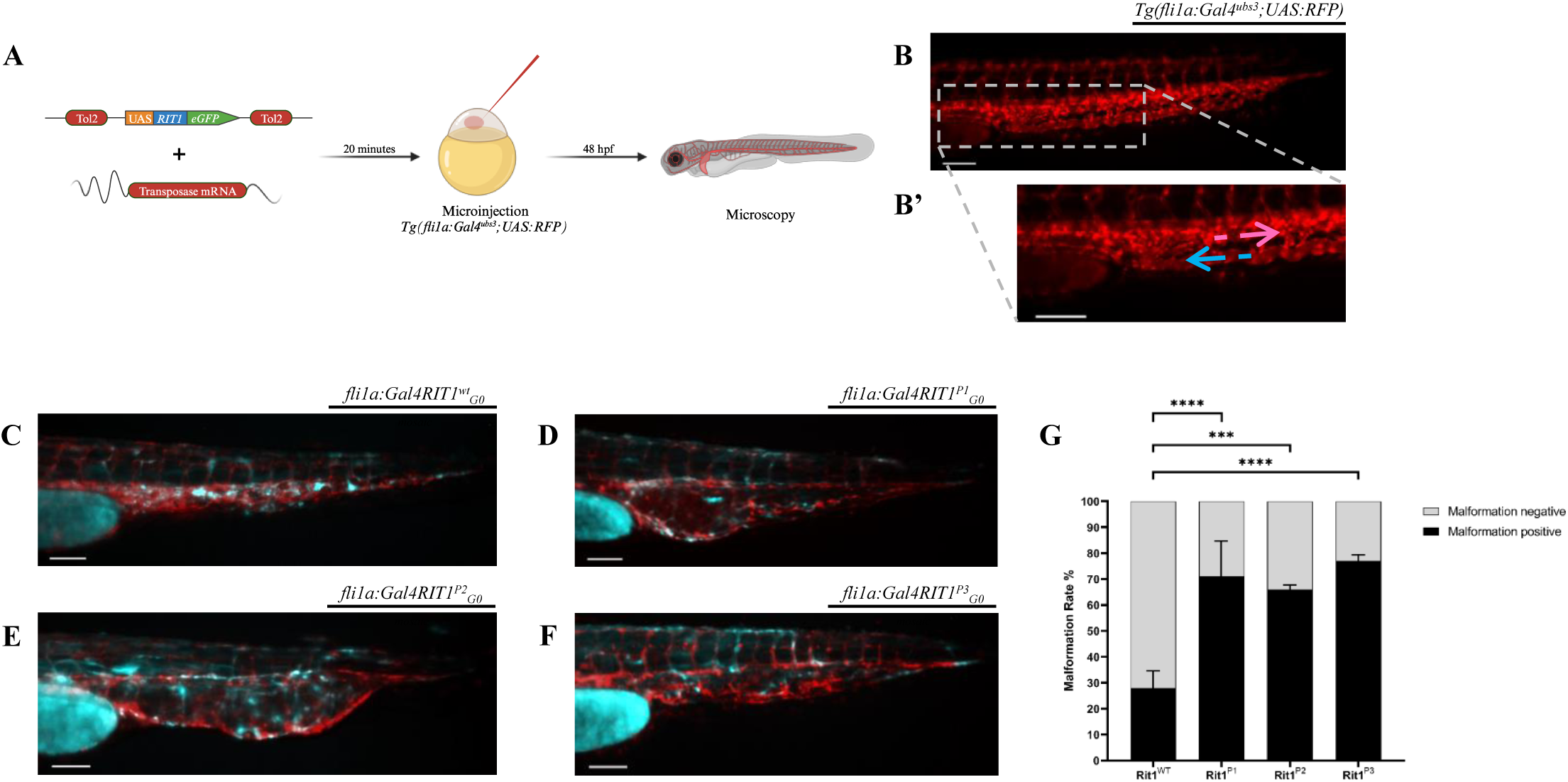
Endothelial-specific mosaic expression of *RIT1* variants leads to the formation of AVM in zebrafish embryos. A. Experimental layout: The plasmid containing human wildtype *RIT1* or *RIT1* variants, under the control of a UAS element and linked to GFP with a P2A sequence is mixed with transposase mRNA and injected into the one-cell stage of *Tg(fli1a:Gal4; UAS:RFP)* embryos. Thereafter, embryos are either examined at 48 hpf or treated with trametinib from 14 to 48 hpf followed by an examination. B. Vascular network (red) in the tail of an uninjected *Tg(fli1a:Gal4; UAS:RFP)* embryo; the dashed white box is enlarged in **B’**; arrows represent the direction of arterial and venous blood flow (pink and blue arrow, respectively). C. Tail of a *fli1a:RIT1^wt^_G0 mosaic_* embryo with normal vasculature (red) despite overexpression of the construct, as indicated by the eGFP expression (cyan). D. Tail of *fli1a:RIT1^P1^_G0 mosaic_* embryos. Scale bar 50 µm. E. Tail of *fli1a:RIT1^P2^_G0 mosaic_* embryos. Scale bar 50 µm. F. Tail of *fli1a:RIT1^P3^_G0 mosaic_* embryos. Note the malformed vasculature with a fusion of the dorsal aorta and the caudal vein as well as dilation of the vessel (in D, E and F). Scale bar 50 µm. G. Quantification of the vascular anatomy at 48 hpf following the injection of plasmids containing the indicated *RIT1* variants, with and without treatment. n =2. Fisher’s exact test, two-tailed. ****P(WT vs P1) = 0.00002, ***P(WT vs P2) = 0.0007, ****P(WT vs P3) = 0.00001. Data are presented as mean ± SEM.

**Fig 4.**
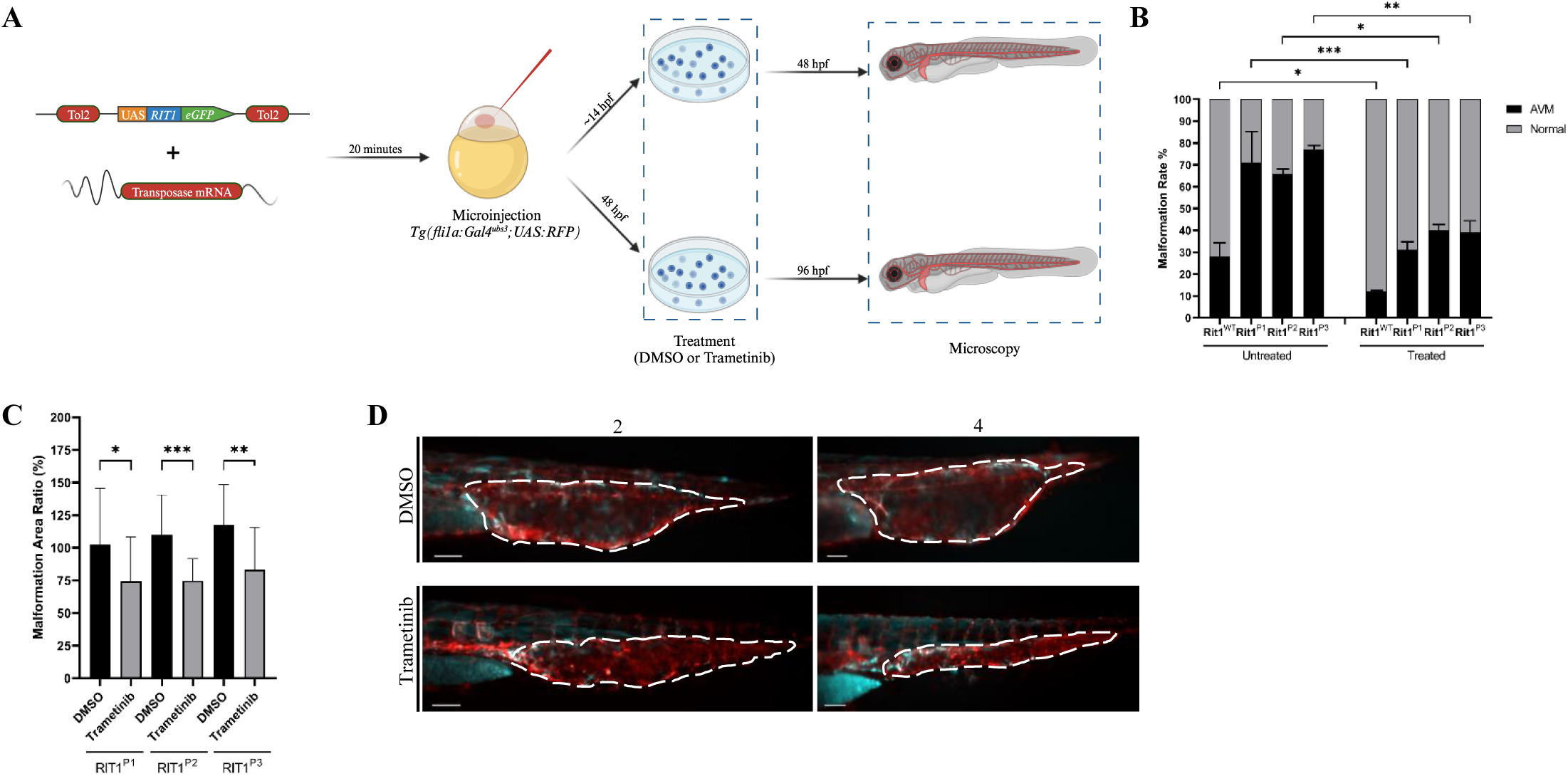
*RIT1* variants injected zebrafish embryos respond to early and late treatment with MEK inhibitor trametinib. A. Experimental plan to assess the effect of trametinib on the RIT1 injected zebrafish embryos after early and late treatments. B. Quantification of the vascular anatomy at 48 hpf following the injection of plasmids containing the indicated RIT1 variants and treatment at 12hpf. n=2. Fisher’s exact test, two-tailed. *P = 0.0405, **P = 0.0030, ***P = 0.0004. Data are presented as mean ± SEM. C. Quantification of the relative change in the size of the AVM-like lesions after 2 days of treatment. n=2. Unpaired t-test, two- tailed. *P = 0.0358, **P= 0083, ***P= 0.0005. Data are presented as mean ± SD. D. Relative change in the size of the AVM-like lesions after 2 days of treatment with DMSO and Trametinib. Treatment started at 2dpf.

### Trametinib induced reduction in AVM size and bleeding frequency in P1

As described above, P1 had a refractory disease with recurrent life-threatening bleeding episodes. Due to the aggressive course of the disease, treatment with thalidomide ^19^ was started but was only transiently effective before symptoms deteriorated again (Fig 5A and B). The MRI showed a large AVM of the right side of the face that was progressive over time (Fig S3). Because of increasing disease severity, off-label treatment with trametinib (0.25 mg per day (1/2 capsules), 0.023 mg/kg/d) was started. Dosage was increased to 0.5 mg per day (1 capsule),

**Fig 5.**
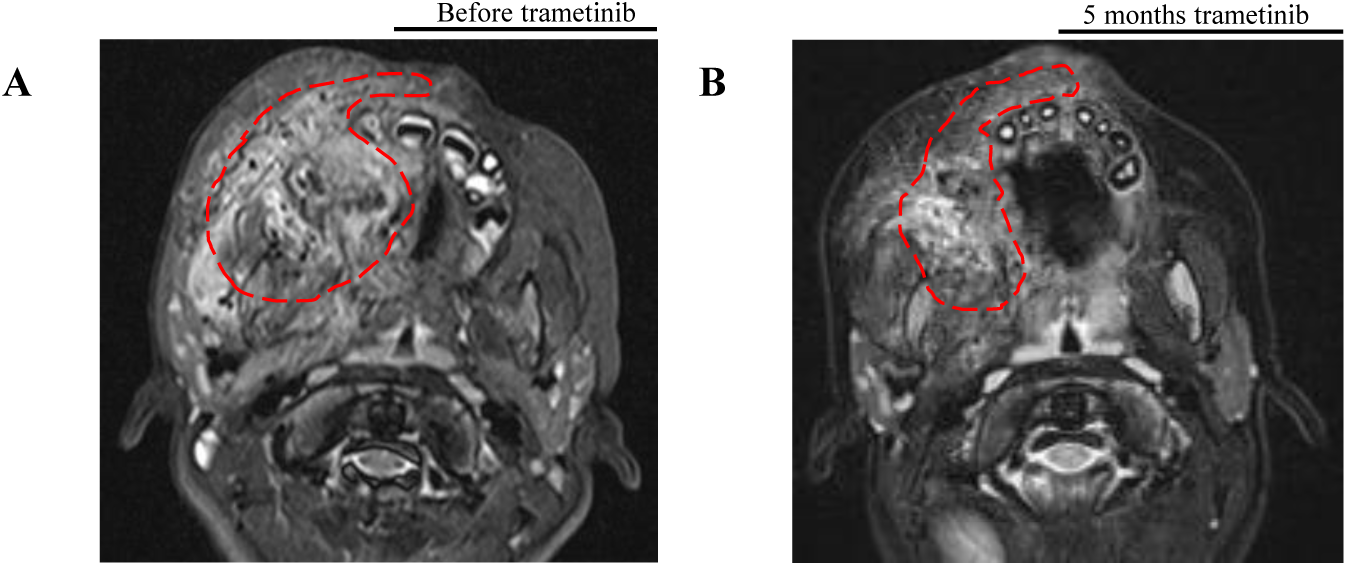
Response to targeted therapy in P1. A. MRI (transversal T2 STIR) of the Patient P1 before the start of trametinib treatment; red dashed line indicates the extent of the AVM. B. MRI (transversal T2 TSE Dixon) of the Patient P1 after 5 months of treatment with trametinib; red dashed line indicates the now smaller extent of the AVM.

0.045 mg/kg body weight after one month. Trametinib led to a significant clinical response with a decrease in frequency and severity of bleeding episodes, a regression of the AVM as observed in the MRI, and a shrinking of the affected cheek. Due to improved disease control, the patient was able to attend preschool for the first time in her life. The patient tolerated trametinib treatment without significant adverse events and remained on this therapy for 9 months. After 4 months of therapy, while on trametinib, the patient developed spontaneous rhinoliquorrhea and was diagnosed with a frontoethmoidal encephalocele, which, retrospectively, was already present in the first MRI at 6 months of age. She then underwent three neurosurgical operations, however the cerebrospinal fluid (CSF) leak persisted and a ventriculoperitoneal (VP) shunt was implanted. After the placement of the VP shunt, the rhinoliquorrhea stopped. The VP shunt was replaced after 3 months due to a defect of the parietal skin overlying the shunt line. One month later, the patient developed a pneumococcal meningitis with cerebral edema and herniation leading to death 4 months after first VP shunt implantation. We hypothesize that the frontoethmoidal encephalocele with difficult dural closure was a predisposing factor for this lethal infection but cannot rule out a contribution of trametinib treatment to this event. However, due to the absence of other adverse events (such as neutropenia or skin toxicity), and the safety profile of trametinib that does not include immunosuppressive effects, we consider this tragic fatal event after 9 months of trametinib treatment as unrelated to this medication.

## Discussion

In this report, we describe three novel somatic activating *RIT1* indel variants in patients with peripheral fast-flow malformations. The exact prevalence of *RIT1* variants as the cause for AVM cannot be determined exactly, but with a total of three cases in the cohorts of vascular anomaly patients studied by this consortium they are obviously much less common than variants of *MAP2K1* or *KRAS*. Nevertheless, we strongly recommend that *RIT1* should be included in panels for genetic testing of patients with vascular malformations.

All three mutations are located close to the switch 2 domain of *RIT1*, a domain that also harbors *RIT1* germline mutations commonly associated with Noonan syndrome. Interestingly, similar indel variants at the switch 2 domain of the RAS GTPases *KRAS* and *HRAS* have repeatedly been described in vascular anomalies^20^. RIT1 indel mutations led to a strong ERK hyperphosphorylation that was much more pronounced than in Noonan-associated RIT1 missense changes. Considering the presumed role of RIT1 in the RAS-MAPK signaling pathway, treatment with a SHP2 inhibitor – as expected – did not influence ERK phosphorylation in cells transfected with the indel variants. In contrast, MEK inhibition completely rescued ERK hyperphosphorylation. These data suggest that RIT1 indel mutations act through overactivation of the canonical RAS-MAPK pathway.

To further assess the impact of the novel *RIT1* variants on vascular development, we used an approach that mimics the endothelial mosaicism that occurs in patients. In our previous work, we assessed various *TEK* mutations by endothelial-specific mosaic expression in zebrafish embryos and observed the development of venous malformations^18^. In the current work, we observed that *RIT1* mutations induced AVM-like lesions in zebrafish embryos, further confirming that overactivation of RAS-MAPK signaling caused by *RIT1* indels has a deleterious effect on vascular development. Moreover, we show that MEK inhibition not only normalized ERK phosphorylation *in vitro* but also restored normal vascular development and decreased AVM-like lesion size *in vivo* during early and late treatment, respectively. These data further validate RAS-MAPK hyperactivation as the major driving mechanism for AVM formation and maintenance in the presence of the novel *RIT1* indel variants.

These findings encouraged us to the off-label use of the cancer drug trametinib, a selective MEK inhibitor, in our severely affected patient P1, who indeed responded very well with a significant reduction of AVM size and associated complaints, such as bleeding episodes. Unfortunately, the patient developed fatal meningitis, most likely due to incidental encephalocele. While we consider this event as not related to trametinib treatment, it further highlights the need for controlled studies in the field of vascular anomalies, to assess treatment efficacy and tolerability and to advance care for patients with these diseases into an era of evidence-based personalized medicine.

In summary, our work introduces *RIT1* as a novel gene implicated in the pathogenesis of AVM. Functional testing *in vitro* and *in vivo* demonstrated the capacity of the novel *RIT1* variants to hyperactivate the RAS-MAPK pathway and induce the development of AVM. MEK inhibition led to biochemical normalization, prevention of AVM formation, as well as decreasing AVM size. We also present the first promising data on the use of trametinib in a patient with a somatic *RIT1* mutation, encouraging further investigation of MEK inhibition in patients with AVM in future clinical trials.

## Materials and Methods

### Patients / Study Approval

All subjects, and/or their legal guardians, gave written informed consent to genetic investigations, which were carried out with approval by the institutional review boards of the University Hospital Regensburg, Germany (17-854-101), University Hospital of Bern, Switzerland (2017-01960), and Boston Children’s Hospital, Boston, MA, USA (IRB- P00025772).

### Genetic testing

*RIT1* was tested in a total of 662 samples by the partners’ laboratories (235 in Magdeburg, 114 in Bern, 313 in Boston), including all types of vascular anomalies.

A tissue biopsy of the AVM of P1 was submitted to the Institute of Human Genetics, University Hospital Magdeburg, and the genomic DNA was extracted. Assuming a mosaic mutation as the cause of the disorder, ultradeep sequencing and enrichment using an Agilent SureSelect XT HS2 Custom Enrichment Panel with molecular barcoding (UMIs, 3bp duplex) (Agilent Technologies) were performed. The library was sequenced on a NextSeq550 instrument (Illumina), 2x150 bp paired-end reads. The target regions had a mean coverage of >3,000x after demultiplexing. The varvis 1.20.0 analysis software (Limbus Medical Technologies GmbH) was used for analysis.

P2 has been included in the Bernese Congenital Vascular Malformation Registry, a prospective cohort of congenital extracranial/extraspinal vascular malformations that have been enrolling consecutive patients since 2008 ^21^. As of October 2020, genetic testing is performed on tissue available from diagnostic biopsies of vascular malformations, using the TruSight Oncology 500 (TSO500; Illumina) Next Generation Sequencing (NGS) gene panel.

Resected tissue from P3 underwent targeted DNA NGS testing via the OncoPanel assay at the Center for Advanced Molecular Diagnostics (CAMD) at Brigham and Women’s Hospital^22^. DNA was isolated using standard extraction methods (QIAGEN) and quantified with PicoGreen-based double-stranded DNA detection (Thermo Fisher Scientific). Indexed sequencing libraries were prepared from 50-ng sonically sheared DNA samples using Illumina TruSeq LT reagents (Illumina). Extracted DNA underwent targeted NGS using the KAPA HTP Library Preparation Kit (Roche), a custom RNA bait set (Agilent SureSelect) and sequenced with the Illumina HiSeq 2500 system.

### Cell culture and Western Blot

Three million HEK293T cells were seeded in 10 cm cell culture plates supplemented with DMEM containing 10% fetal bovine serum (FBS) 12 hours prior to transfection. At around 70% confluency levels cells were transfected using TurboFect transfection reagent (Thermo Fisher #R0532), with Flag-tagged RIT1 variants in pCDNA constructs or empty vector (EV) as the negative control. The medium was refreshed the next day and at 48 hours post-transfection, cells were washed in ice-cold phosphate-buffered saline (PBS) and lysed in ice-cold lysis buffer, containing 50 mM Tris/HCl pH 7.5, 5 mM MgCl2, 100 mM NaCl, 1% Igepal CA-630, 10% glycerol, 20 mM ß-glycerolphosphate, 1 mM Na-orthovanadate, EDTA-free inhibitor cocktail 1 tablet/50 ml. After the addition of Laemmli sample buffer, the samples were subjected to SDS-PAGE (12.5% polyacrylamide). Blots were detected by immunoblotting using a mouse anti-γ-Tubulin antibody (Sigma #T5326), a mouse anti-FLAG antibody (Sigma #F3165), a rabbit anti-ERK antibody (Cell signaling technology #9102), and a rabbit anti-p- ERK antibody (Cell signaling technology #4370), respectively. The immunoblots were detected using an Odyssey Fc Imaging System (LI-CORE Biosciences) and analyzed by Image Studio Lite Ver 5.2.

### Zebrafish husbandry

Maintenance and breeding of zebrafish (*Danio rerio*) were performed in the fish facility of the Developmental Biology, Institute for Biology I, University Freiburg under standard conditions. Only embryos up to 5 days post-fertilization were used. All experiments were carried out in accordance with German laws for animal care and the Regierungspräsidium Freiburg.

### Plasmid preparation

Plasmids were designed using ApE—A plasmid editor version 3.0.8. *Homo sapiens RIT1* sequence was obtained from the online database Ensembl (Transcript ID: ENST00000368323.8), minimally codon optimized for *Danio rerio* and ordered as a plasmid including Tol2 sites, a UAS promoter, RIT1^P2^, and P2A-GFP from Twist Bioscience (South San Francisco, CA, USA). Plasmids were purified using Wizard Plus SC Minipreps DNA Purification Systems (Promega, Walldorf, Germany, A1330) according to the manufacturer’s instructions.

### Mutagenesis

*RIT1^wt^* as well as all other *RIT1* mutations analyzed in this study were derived from the UAS:RIT1^P2^-P2A-GFP construct using Q5 Site-Directed Mutagenesis (New England Biolabs, E0554S). Corresponding mutagenesis primers were designed using NEBaseChanger version 1.3.3. All plasmids were sequenced to confirm the expected sequence.

### Tol2 transposase mRNA Synthesis

8 µg of the plasmid that contains the transposase gene under control of the SP6 promoter were linearized using 4 µl of NotI-HF enzyme (New England Biolabs) for 1 hour at 37°C. The digested sample was purified using the QIAquick PCR Purification Kit according to the manufacturer’s protocol. Capped Tol2 transposase mRNA was synthesized from purified DNA using the mMESSAGE mMACHINE™ SP6 Transcription Kit (ThermoFisher Scientific) according to the manufacturer’s protocol. The resulting mRNA was separated into 5 µl aliquots and stored at –20°C to prevent freeze-thaw cycles.

### Plasmid injection

The construct was then injected into *Tg*(*fli1a:Gal4FF^ubs^*^3^*; UAS:RFP)* embryos at the one-cell stage together with Tol2 transposase mRNA ^23^, both at a concentration of 30 ng/µl. For better readability, *Tg*(*fli1a:Gal4FF^ubs^*^3^*; UAS:RFP)* embryos injected with a gene of interest (e.g. UAS:RIT1^P^^1^-P2A-GFP) are abbreviated as *fli1a:RIT1^P1^_G0mosaic_* instead of *Tg*(*fli1a:Gal4FF^ubs^*^3^*; UAS:RFP)* and *Tg(UAS:RIT1^P^*^1^*-P2A-GFP)_G0mosaic_*.

### Fluorescence microscopy

An Axio Examiner D.1 fixed stage fluorescence microscope (Carl Zeiss) was used to acquire images. This microscope has a Zeiss AxiCam MRm camera for image acquisition, which has a high resolution and sensitivity for capturing images. The microscope is equipped with 5x objective lenses and a 10x/23 eyepiece. Final images/videos have a magnification of 50x. The numerical aperture of the objective lenses is 0.15, which provides a good balance between resolution and depth of field. During the imaging acquisition, the sample was placed in 3 ml of E3 medium with tricaine (0.168 mg/ml) at room temperature. The sample had two fluorochromes, RFP and eGFP, to visualize different aspects of the sample. Image and video acquisition was performed using ZEN Pro software (Carl Zeiss). After the image/video has been acquired, it was processed using ZEN 3.5 Lite Blue edition (Carl Zeiss) software. The color cyan is assigned for RFP channel and magenta is assigned for eGFP channel. Images and videos were exported as .png and .AVI (uncompressed) files, respectively.

### Pharmacological treatments

For pharmacological treatments, injected zebrafish embryos were randomized into control and treatment groups. From the 10-somite stage or from 48 hpf on, embryos of the treatment group were transferred in E3 Medium (5 mM NaCl, 0.17 mM KCl, 0.33 mM CaCl_2_, 0.33 mM Mg_2_SO_4_) containing 0.2 mM 1-phenyl 2-thiourea (PTU; Sigma, Taufkirchen, Germany, P7629) and MEK1/2 inhibitor Trametinib (MedChemExpress, GSK1120212; 10 mg) using 100× stock solutions dissolved in dimethyl sulfoxide (DMSO; Sigma, D2650). The treatment dose of trametinib was chosen at 100 nM, according to our previous publication ^18^, and by repeating of the toxicity assay in zebrafish embryos. Embryos of the control group were raised in E3 medium with 0.2 mM PTU and DMSO (equal amount to the treatment group). The response of the AVM-like lesion size to trametinib was calculated as follows: each embryo with an AVM-like lesion was imaged at the Axio Examiner, and the area of the lesion was divided by the area of the entire embryo to give a relative area of the malformation at 2 dpf. This was done to control for different embryo sizes and different embryo growth rates. This measurement was then repeated at 4 dpf (after treatment) and the relative area of the AVM-like lesion at 4 dpf was divided by the relative area 2 dpf, followed by multiplication with 100 to give a result in percent. A result greater than 100% showed an AVM-like lesion growing in size in relation to the embryo, a result less than 100% showed a regressing lesion.

### Statistical analysis

The statistical analysis was performed using two-tailed Fisher’s exact test of significance for malformation rate analysis and early pharmacological treatments and two-tailed unpaired t-test for late pharmacological treatment experiments and one-tailed Student’s t-test for *in vitro* experiments in GraphPad Prism version 9.0. The legends of the figures include information on sample sizes and significance. P-value of < 0.05 was regarded as significant. Analyzed data for zebrafish malformation rates and early pharmacological treatments was obtained from at least two independent experiments for each variant. The number of injected embryos ranged from 51 to 102 for each variant. Data are presented as mean ± SEM for malformation rate calculations and early pharmacological treatments and mean ± SD for late pharmacological treatments. P- value ∗∗ < 0.01; ∗∗∗ < 0.001; ∗∗∗∗ < 0.0001; for P-values < 0.05 and > 0.01 the actual value is reported.

## Data availability

The data that support the findings of this study are available from the corresponding author upon reasonable request.

## Acknowledgments

The authors first and foremost acknowledge the patients and their families for providing their information and allowing us to conduct this work. We further acknowledge the Center Vascular Anomalies at the Freiburg Center for Rare Diseases, and the Hilda Biobank at the Department of Pediatrics and Adolescent Medicine, Freiburg, Germany for their contribution. Four of the authors of this publication are members of the Vascular Anomalies Working Group (VASCA WG) of the European Reference Network for Rare Multisystemic Vascular Diseases (VASCERN)—Project ID: 769036. Five of the authors are members of the German Reference Network for Vascular Anomalies. MZ and MRA received funding from the German Federal Ministry of Education and Research (BMBF): German Network of RASopathy Research (GeNeRARe; FKZ 01GM1902A and 01GM1902C). NM was part of the International Master Program in Biomedical Sciences (IMBS) and acknowledges the support received throughout the program. FB and MRA received funding from the German Research Foundation (Grant numbers: DFG AH 92/8-3 and IRTG 1902-p6) and the Foundation for Ageing Research of the HHU Düsseldorf. AH received the Walter Benjamin Fellowship from the German Research Foundation (Project number: 458322953). IB (Principle Investigator) and JR (Co-Investigator) were supported by the Swiss national Research Foundation, Synergia grant CRSII5_193694 2021-2024.

## Contributions

**Friedrich G. Kapp**: clinical care of patient, conceptualization of the study, writing - original draft preparation, writing - review and editing. **Farhad Bazgir**: *in vitro* analyses, formal analysis. **Nagi Mahamammadzade**: *in vivo* analyses, formal analysis. **Axel Karow**, **Caroline Seebauer**, **Walter A. Wohlgemuth**: clinical care of patients, **Annegret Holm**, **Whitney Eng**: clinical care of patients, original draft preparation, writing – review and editing. **Whitney Eng**, **Denny Schanze**, and **Martin Zenker**: genetic analyses. **Pia Kröning**: writing - original draft preparation. **Charlotte M. Niemeyer**, **Yvonne Döring**, **Mohammad R. Ahmadian**: resources. **Iris Baumgartner** and **Jochen Rössler**: conceptualization, writing - original draft preparation, writing - review and editing, resources. All authors provided intellectual input, critical feedback, discussed results, and approved the final manuscript. Authors **Friedrich G. Kapp**, **Farhad Bazgir**, **Nagi Mahammadzade**, **Iris Baumgartner** and **Jochen Rössler** contributed equally to this work.

## Abbreviations

AVM: arteriovenous malformation
CSF: cerebrospinal fluid
dpf: days post fertilization
hpf: hours post fertilization
ISSVA: International Society for the Study of Vascular Anomalies
MRI: magnetic resonance imaging
NGS: next generation sequencing
VP: ventriculoperitoneal

