## Supplemental Figure S1 for "Somatic RIT1 indels identified in arteriovenous malformations hyperactivate RAS-MAPK signaling and are amenable to MEK inhibition"

**Supplemental Figure S1. MR angiography of P1 and P2 in AVM tissue.**

A. The MR angiography of the P1 shows increased perfusion on the right side of the face (left side of the MRI angiography).
B. MR angiography of the P2 shows the large AVM connected to the subclavian and the thyrocervical trunk with multi-chambered central nidus.

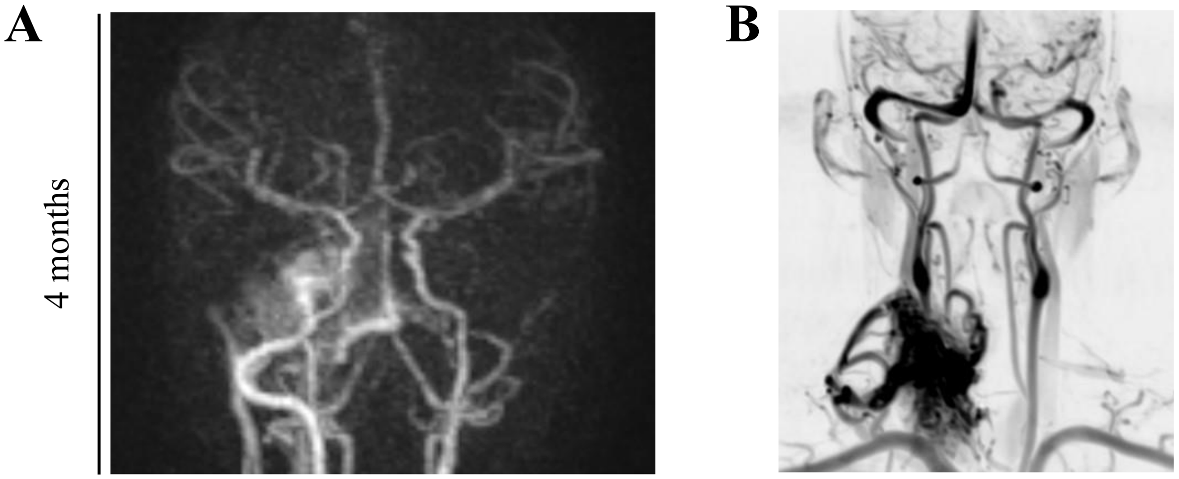
