## Supplemental Figure S2 for "Somatic RIT1 indels identified in arteriovenous malformations hyperactivate RAS-MAPK signaling and are amenable to MEK inhibition"

**Supplemental Figure S2. Schematic of the RAS signaling pathway.**

SHP2 is upstream, MEK further downstream in the RAS-MAPK signaling pathway; created with BioRender.com.

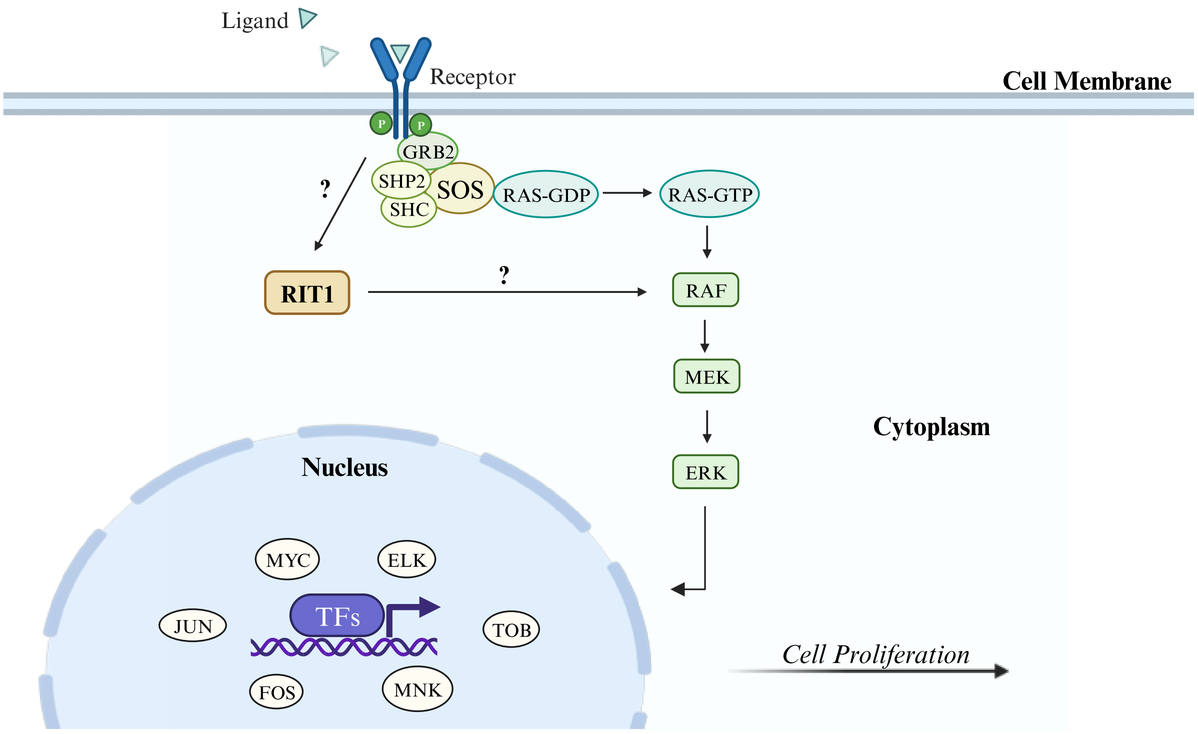
