## Supplemental Figure S3 for "Somatic RIT1 indels identified in arteriovenous malformations hyperactivate RAS-MAPK signaling and are amenable to MEK inhibition"

**Supplemental Figure S3. Patient P1 – Radiological evolution of disease**.

**(Upper panels)** MRI images show the AVM's progression over time, non-response to thalidomide, and regression under trametinib treatment.
**(Lower panel)** Image of an angiography showing the extent of the Onyx cast on the right side of the face.

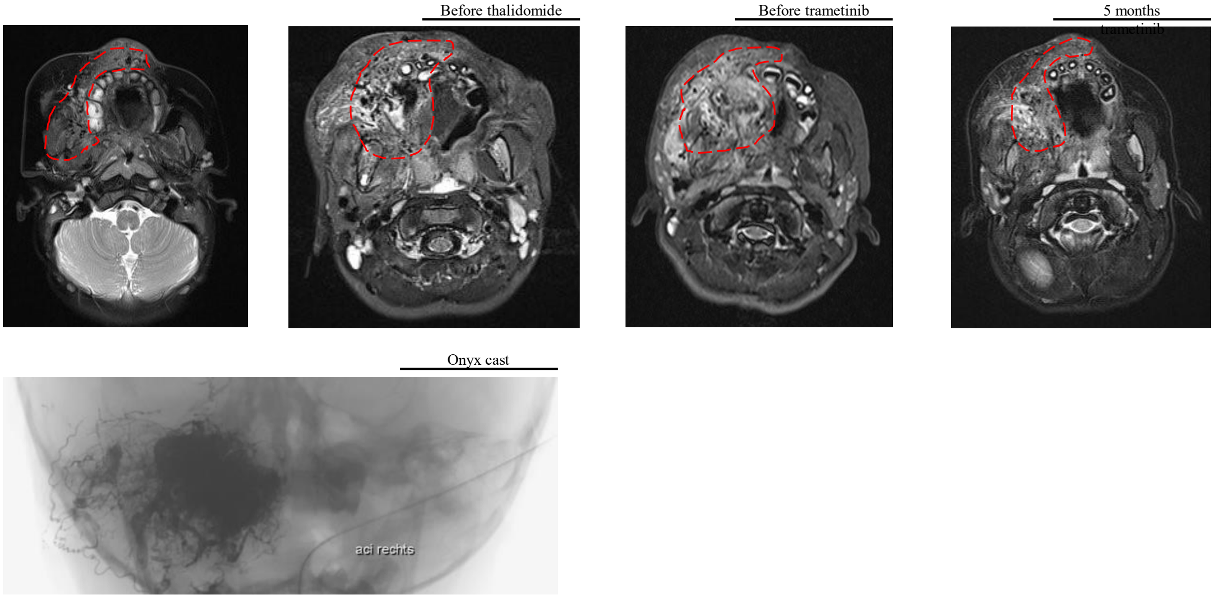
