## Supplemental Video S1 for "Somatic RIT1 indels identified in arteriovenous malformations hyperactivate RAS-MAPK signaling and are amenable to MEK inhibition"

**Supplemental Video S1. Comparison of normal circulation and Aberrant connection of aorta and caudal vein with fusion and dilation of vasculature in the tail distal to the AVM.**

A. Notice that the blood in the dorsal aorta of the uninjected *Tg(fli1a:Gal4)* fish flows to the end of the tail and then returns in the caudal vein. Scale bar 50 µm. Link to the video.
B. Notice the aberrant flow in the dorsal aorta of the $RIT1^{P2}$ injected fish which moves into the caudal vein proximal in the tail, as well as the dilation of distal vessels. Scale bar 50 µm. Link to the video.
